# Auditory steady-state response deficits in Fragile X Syndrome implicate deficits in stimulus representation maintenance and GABAergic modulation

**DOI:** 10.1101/2025.01.29.25321365

**Authors:** Jordan E. Norris, Lisa A. De Stefano, Walker S. McKinney, Lauren M. Schmitt, Makoto Miyakoshi, Christina Gross, Sebastian Piloto, Braeden Heald, Ernest V. Pedapati, Craig A. Erickson, John A. Sweeney, Lauren E. Ethridge

## Abstract

**Background:** Fragile X Syndrome (FXS) is a rare, neurodevelopmental disorder caused by a mutation to the Fragile X messenger ribonucleoprotein 1 (*Fmr1*) gene and characterized by sensory processing abnormalities and sensitivities, including neural auditory oscillatory disruptions and reduced neural entrainment to chirp stimuli. The present study aims to evaluate the 40 Hz auditory steady state response (ASSR) in FXS to evaluate stimulus representation maintenance in FXS.

**Methods:** Adolescents and adults (N = 67; 34 FXS and 33 age, sex-matched typically developed controls (TDC)) completed a 40 Hz auditory steady state task during electroencephalography (EEG). Time-frequency analyses using Morlet wavelets were completed to evaluate intertrial phase coherence (ITC) and event-related spectral perturbation (ERSP), including characterization of the transient and sustained components of the 40 Hz ASSR.

**Results:** Both ITC (*p* = .003) and ERSP (*p* = .004) at 40 Hz were reduced for FXS compared to TDC. Interestingly, TDC exhibited a significantly elevated early, transient component (100 – 400 ms) which reduced in both ITC and ERSP during transition to the sustained component (650 – 3000 ms) whereas FXS were consistently reduced across the ASSR suggesting a reduced ability for FXS to mount a transient response.

**Conclusions:** Individuals with FXS exhibit robust reductions in magnitude and temporal precision of neural entrainment to the steady state stimulus. The reduced ability to mount a transient response may represent reduced GABAergic modulation where the overall reduction in ITC and ERSP may reflect reduced excitatory/inhibitory balance between NMDA and GABAergic input.

## Background

Fragile X Syndrome (FXS) is a rare, neurodevelopmental disorder caused by a trinucleotide repeat expansion ≥ 200 CGG copies in the 5’ untranslated region of the Fragile X messenger ribonucleoprotein 1 (*Fmr1*) gene leading to gene methylation, and loss of the protein product, Fragile X messenger ribonucleoprotein (FMRP) (Hagerman et al.,2017; Salcedo-Arellano et al., 2020). Loss of FMRP results in overproduction of synaptic-associated proteins, increased dendritic spine density, and immature dendritic spine morphology which impacts neural communication efficiency (Hagerman et al., 2017; Huebschman et al., 2020; Pfeiffer & Huber, 2009; Salcedo-Arellano et al., 2020). The loss of efficiency is particularly detrimental to auditory processing which requires precise and fast synapses to carry out the unique functions of the auditory system where quick and precise temporal integration is largely a product of gamma band (> 30 Hz) oscillations (McCullagh et al., 2020; Metzner & Steuber, 2021). Auditory hyperexcitability is a robust and well-documented phenotype in FXS rodent models, which support a broader frequency tuning curve of individual auditory neurons, and abnormal spectrotemporal processing, which negatively impact auditory processing efficiency (Rotschafer & Razak, 2013, 2014; McCullagh et al., 2020).

Increased auditory processing abnormalities, documented clinically and via electroencephalography (EEG), and clinical auditory sensory hypersensitivities are common in FXS (Rais et al., 2018; Ethridge et al., 2017; 2019. Previous studies investigating auditory processing in individuals with FXS using EEG utilized the auditory chirp stimulus which consists of broadband noise amplitude modulated by a linearly increasing set of frequencies and thus models temporal precision and flexible processing in the gamma range to a rapidly changing stimulus. Findings from FXS chirp studies identified neural entrainment deficits in the low gamma range (31 – 57 Hz) and broadband increases in gamma power both suggestive of neural hyperexcitability and reduced temporal fidelity in stimulus response, findings also documented in the *Fmr1^-/-^* mouse (Ethridge et al., 2017; 2019; Jonak et al., 2020). Reduced low gamma ITC and elevated gamma power were linked to increased features of autism, elevated sensory issues, and reduced IQ making the chirp a good FXS biomarker when clinically targeting sensory-related symptoms. Auditory steady state response (ASSR) stimuli, which are modulated by a single frequency over a longer period of time, by contrast, allow for more in-depth analysis of neural processing dynamics and maintenance of stimulus fidelity over time at that target frequency. This study represents the first evaluation of the ASSR in human participants with FXS and provides an in-depth evaluation of low gamma responses in FXS.

The increased use of the auditory evoked steady state responses as a measure of brain disruptions in neuropsychiatric conditions has generated a robust literature detailing mechanisms contributing to the generation and maintenance of the ASSR, of which 40 Hz has the strongest effect in humans with good test re-test reliability (McFadden et al., 2014). Neuropsychiatric conditions with notable disruptions to the 40 Hz ASSR include schizophrenia, bipolar disorder, and autism, though accounts of ASSR disruptions in autism are less consistent (Arutiunian et al., 2023; Grent-‘t-Jong et al., 2023; Ono et al., 2020; Seymour et al., 2020; Wilson et al., 2006). Interestingly, ASSR abnormalities in schizophrenia are linked to reduced cognitive performance on memory and attention tasks suggesting the ASSR may serve as a general biomarker of gamma-range activity disruptions (Koshiyama et al., 2024; Parciauskaite et al., 2020). Further, gamma range ASSR is linked to cognitive flexibility in typically developed individuals providing further evidence in support of a link between ASSR and cognitive-related gamma activity in addition to sensory processing disruptions (Parciauskaite et al., 2020).

The use of the 40 Hz steady state paradigm provides unique insight into the fidelity of auditory entrainment which may provide further information about mechanistic disruptions and how they contribute to sensory-related deficits in FXS. The ASSR is a robust index of auditory processing and likely represents a marker of inhibitory mechanisms (Grent-‘t-Jong et al., 2021; Sugiyama et al., 2021). Studies in neuropsychiatric conditions and pharmacological evidence identified both gamma-aminobutyric acid (GABA) and N-methyl-D-aspartic acid (NMDA) receptor contributions to the ASSR. Specifically, NMDA receptor (NMDAr) action on parvalbumin positive (PV+) interneurons is necessary for generating the ASSR and GABAergic input modulates the response (Kozono et al., 2019; Sivarao et al., 2013, 2016). Ultimately, inappropriate NMDAr activity may dysregulate fast-spiking GABAergic interneurons and bias the excitatory/inhibitory balance in favor of excitation (Hashimoto et al., 2003; Tada et al., 2013).

In rats with concurrent scalp-level EEG and multi-channel local field potential recordings in primary auditory cortex (A1), increased 40 Hz ASSR from scalp EEG corresponds to increased temporal consistency or reduced latency variability across A1 layers suggesting better phase coherence is a byproduct of increased temporal precision (Johnson et al., 2024). Recent steady state investigations in *Fmr1* knockout (*Fmr1^-/-^*) mice and rats demonstrated attenuated intertrial phase coherence (ITC) and event related spectral perturbation (ERSP) to 40 Hz auditory steady state stimuli (Jonak et al. 2024; Kozono et al., 2020). Further, the ASSR in *Fmr1^-/-^* mice demonstrated comparable results to the auditory chirp highlighting the viability of the ASSR as a potential clinical target which builds on previous auditory chirp findings. Evaluating the ASSR in FXS humans may provide increased mechanistic insight into the molecular disruptions underlying reduced auditory neural entrainment in the low gamma range and provide a robust biomarker for translation of pharmacological interventions from preclinical models to clinical trials.

The current study utilized the auditory steady state paradigm to evaluate the 40 Hz ASSR in adults and adolescents with FXS. We hypothesized that the steady state paradigm would capture a comparable phenotype to that captured by the auditory chirp paradigm. Specifically, we predict reduced ITC in the low gamma range in FXS relative to typically developing controls (TDC),that would be consistent across the duration of the ASSR. We also conducted an exploratory investigation of the 40 Hz response to evaluate potential novel mechanisms indexed by the steady state paradigm and evaluated its potential relationship with peripheral FMRP levels. Given the high test-retest reliability, documented ASSR phenotype in *Fmr1^-/-^* mice, and literature documenting molecular mechanisms, the ASSR may allow for further insight into the FXS auditory processing allowing for the establishment of ASSR as a biomarker for translational purposes.

## Methods

### Participants

Participants were 67 adolescents and adults (FXS: N = 34, 12 females, mean age = 30.90, SD = 9.60; age and sex matched TDC: N = 33, 11 female, mean age = 27.34, SD = 7.70) aged between 13 – 50.25 (M = 29.15, SD = 8.83). Of the 67 participants, 64 were white (94%), 1 was Black (1.2%), 2 were Asian (3%), and 1 identified as other race (1.2%). Participant ethnicity was largely non-Hispanic (65; 97%). FXS status was confirmed using Southern Blot/Polymerase Chain Reaction analysis. Individuals with a history of seizures were excluded. As with previous electrophysiological evaluations in FXS, individuals were not excluded for the presence of commonly prescribed psychiatric medications because doing so would result in a non-representative sample of the FXS population (see supplement for details). However, the current study included individuals on anti-convulsant medications for mood regulation, unlike in past studies. We confirmed that all EEG outcomes for the individuals on anti-convulsant medications were within 1 standard deviation from the mean for inclusion (Supplemental Table 1).

Caregivers of FXS participants provided clinical information about their child’s behavior via: the Adolescent and Adult Sensory Profile (A/ASP; Brown et al., 2001), the Child Sensory Profile (CSP; Dunn, 2014), the Social and Communication Questionnaire (SCQ; Rutter et al., 2003), the Anxiety Depression and Mood Scale (ADAMS; Esbensen et al., 2003), and the Aberrant Behavior Checklist-Community (ABC-C; optimized for FXS, Sansone et al., 2012). Clinicians or clinical research coordinators administered the Woodcock-Johnson III Tests of Cognitive Abilities Auditory Attention subscale (WJ-III; McGrew and Woodcock, 2001), the Comprehensive Interview Form of the Vineland Adaptive Behavior Scales, Third Edition (Vineland-3; Sparrow et al., 2016), the computerized Test of Attentional Performance for Children (KiTAP; Knox et al., 2012), and the Stanford Binet Intelligence Scale, Fifth Edition (SB-5; Roid, 2003). SB-5 deviation scores were computed for Full-Scale, Abbreviated, Verbal, and Non-Verbal IQ using previously reported methods (Sansone et al., 2014) to minimize SB-5 floor effects common in participants with FXS. Recent work identified a downward shifted but near normal distribution of deviation IQ scores in FXS, highlighting the utility of using deviation IQ scores to capture meaningful variation in FXS (Schmitt et al., 2024).

### FMRP Collection and Analysis

Blood samples were collected from participants then immediately blood spotted and submitted to an immunoassay optimized to detect trace FMRP levels at CCHMC (Boggs et al., 2022). All FMRP levels included in the current study were analyzed within 1 year of blood sample collection (M = .46 years, SD = .20 years, 26-303 days) and 2.07 years (M = .57 years, SD = .44 years, 27-755 days) of the EEG recording.

### EEG Procedure – Steady State Task

The steady state stimuli were 3000 ms long 40 Hz click trains at 100% modulation depth followed by an interstimulus interval randomly jittered between 1500 - 2000 ms. Stimuli were delivered using Sony headphones at 65 dBL SPL via Presentation software while participants passively listened and watched a silent video for both comfort and compliance, similarly to previous studies (Ethridge et al., 2016, 2017, 2019).

### EEG Recording & Analysis

EEG was continuously recorded and digitized at 1000 Hz, filtered from 0.01 to 200 Hz, referenced to Cz, and amplified 10,000x using a 128-channel saline-based Electrical Geodesics system (EGI, Eugene, Oregon) with sensors placed approximately according to the International 10/10 system (Chatrian, 1985; Luu & Ferree, 2005). Data were visually inspected and submitted to artifact removal procedures prior to analysis according to Ethridge et al. (2019). Data were digitally filtered from 0.5 to 120 Hz (12 and 24 db/octave roll-off, respectively; zero-phase; 60 Hz notch: 57 – 63 Hz with harmonics removed up to the Nyquist frequency of the sampling rate), bad channels or channels containing excessive artifact were interpolated (no more than 5%), and segments containing high amplitude artifact or excessive muscle were manually rejected. Data were then submitted to independent components analysis (ICA) via EEGLAB for removal of components containing muscle-related artifacts (e.g., heart rate, eye blinks, etc.) and line noise (Delorme and Makeig, 2004). Data were then referenced to the average of all channels and segmented into 4250 ms epochs (i.e., trials; -500 to 3750 ms). Any epoch containing residual artifact exceeding +/- 150 µV was automatically rejected. The number of valid trials retained after artifact correction was higher for TDC compared to FXS (FXS ranked sums *M* = 27.06 ; Control ranked sums *M* = 41.15; *U* = 325.00, *z* = -2.96 , *p* = .003, r = -.36), therefore trial count was evaluated as a covariate where appropriate and retained when significant.

A final set of 26 channels were selected for analysis. The selection included the 23 sensors distributed over the fronto-central scalp region that were selected *a priori* consistent with our prior work and previous literature capturing activity from auditory cortex (Figure 1; Ethridge et al., 2019; 2017; Luck et al., 2014). An additional 4 channels were added after evaluating the spatial scalp distribution of the steady state signal in our sample and seeing evidence of a more diffuse signal relative to previous auditory responses (Ethridge et al., 2017; 2019).

**Figure 1.**
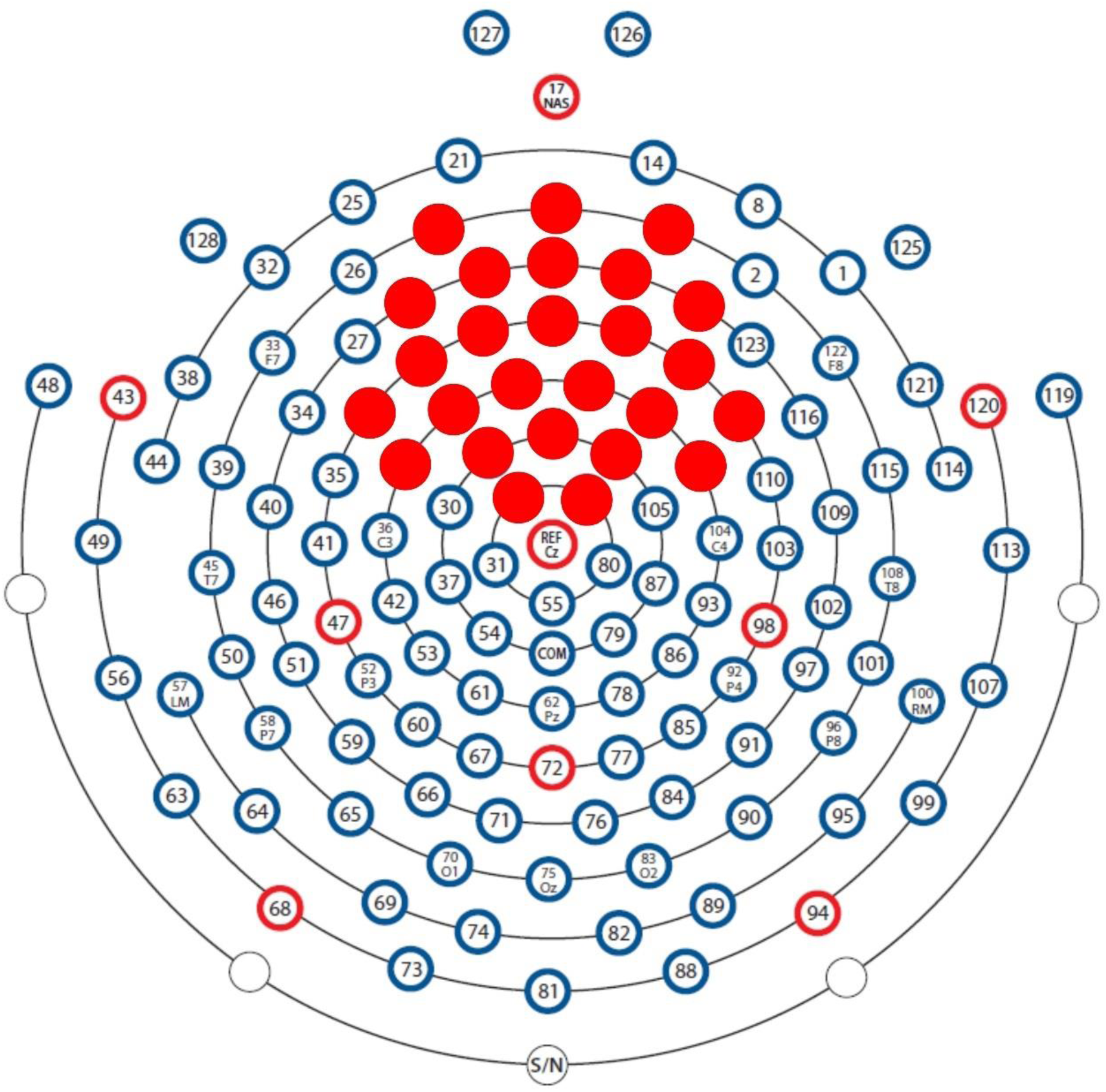
Channel map for the EGI 128 channel net with the 26-electrode selection highlighted in red. Selection was determined by utilizing the same 23-electrode selection from Ethridge et al. (2019) that they based on the N1 component of the event related potential signal from auditory cortices with additional channels added for the current study to capture the more frontal distribution of the ASSR.

The un-baseline corrected trials were analyzed in a time-frequency analysis using Morlet wavelets with a linearly increasing cycle length from 1 to 30 Hz which resulted in roughly 1 Hz steps from the lowest frequency (∼2 Hz) to the highest frequency (110 Hz). Intertrial phase coherence (ITC) measures were calculated to evaluate the degree of phase-locking to the steady state stimulus across trials on average and Event Related Spectral Perturbation (ERSP) was calculated to evaluate absolute power differences. Both ITC and ERSP were obtained using EEGLAB (Delorme and Makeig, 2004). Time frequency values were down sampled to 250 time bins for wavelet calculation. Raw ITC values were corrected for trial count by subtracting the critical r value for each subject based on the remaining clean trials prior to computing ITC-based values (Ethridge et al., 2019). Baseline correction (−500 – 0 ms) was applied to measure ERSP change from baseline at 40 Hz as an exploratory analysis. All frequency and time ranges were selected *a priori* based on methods from Ethridge et al. (2019) and refined to capture the most appropriate measurement window.

The neural response to the 40 Hz stimulus (ITC, ERSP) was first evaluated for the 40 Hz ASSR (35 – 45 Hz). Visualization of the time-frequency plots suggested group differences in spread of activation to this response, so 40 Hz ITC was also comparatively evaluated for a tightened 40 Hz range (i.e., narrow; ∼39 – 41 Hz). Research in schizophrenia and typically developing individuals has shown differences in ITC and ERSP drop-off after stimulus offset, sustained and transient responses to steady state stimuli, and maintenance of steady state response over the course of stimulation (Ethridge et al., 2011; Toso et al., 2024) which reflect different neural mechanisms. To provide a comprehensive view of ASSR in FXS, we also analyzed 40 Hz ITC after stimulus offset, the 40Hz transient (100 – 400 ms) and 40 Hz sustained components (650 – 3000 ms; 35 – 45 Hz), and the 40 Hz response (35 – 45 Hz) in time limited bins (∼75 – 100 ms bins; *M* = 85.17, *SD* = 7.27). We also evaluated the transient and sustained components in the narrowed 40 Hz range. Lastly, we evaluated the 40 Hz response (35 – 45 Hz) between hemispheres by evaluating 40 Hz ITC at channel F3 (i.e., left) and F4 (i.e., right) to replicate analyses on the chirp response in FXS in Norris et al. (2022). Other ITC measures included the 80 Hz harmonic response (75 – 85 Hz) and both stimulus onset (3 – 13 Hz) and stimulus offset responses (3 – 13 Hz). The ERSP measures were not baseline corrected and included theta (4 – 7 HZ), alpha (8 – 13 Hz), and beta (14 – 30 Hz) power. Un-baseline-corrected gamma ERSP was not evaluated because of the sustained entrainment in the gamma range. However, baseline corrected ERSP in the 40 Hz range (35 – 45 Hz) was computed to evaluate the power and the transient (100 – 400 ms) and sustained (650 – 3000 ms) components of the ASSR.

The event related potential (ERP) to both the onset and offset of the stimulus were evaluated using ERPLAB (Lopez-Calderon & Luck, 2014). For the onset, the P1 component (20 – 70 ms), the N1 component (70 – 150 ms), and the P2 component (150 – 320 ms) were computed. Only the N1 (3070 – 3150 ms) and P2 (3150 – 3250 ms) components were computed for the offset because the P1 component was biased for FXS by the steady state response offset.

## Statistical Analysis

All variables were evaluated and tested for normality. For variables that met assumptions of normality, independent samples t-tests were utilized to evaluate group differences with effect sizes reported as Cohen’s d. Variables that failed to meet assumptions of normality were analyzed using independent samples Mann-Whitney U tests with effect sizes reported as r values. Six repeated measures analysis of variance (rmANOVA) tests were used for the exploratory 40 Hz analyses. For all ANOVA results, we report effect sizes as partial eta squared (ES) and observed power (OP). Trial count was included as a covariate where appropriate. Statistical analyses were conducted using IBM SPSS Statistics 29 (IBM Corp, 2023).

### Clinical and EEG Correlations

Exploratory clinical correlations were evaluated between all EEG variables of interest, the clinical assessment battery, and FMRP levels. Additionally, correlations between EEG variables were also evaluated. All exploratory correlations were computed using Spearman’s rho and were considered hypothesis generating, thus no correction for multiple comparisons was applied.

## Results

### ITC in the 40 Hz Range

Independent samples t-tests identified reduced 40 Hz ITC to the steady state stimulus for FXS (*M* = .25, *SD* = .10) compared to TDC (*M* = .33, *SD* = .13), t(65) = 3.06, p = .003, d = .75 (Figure 2). The addition of sex via univariate ANOVA reduced statistical power for the main effect of group (*F*(1, 63) = 28.84, *p* =.12, ES = .97, OP = .33) and the effect of sex (*F*(1, 63) = 1.64, *p* =.42, ES = .62, OP = .09) was not significant. We also evaluated hemisphere differences at electrodes F3/F4, as hemisphere differences were identified in the *Fmr1^-/-^*mice, using a rmANOVA (Jonak et al., 2024). The main effect of group was significant, where the single channel measure of ITC in the 40 Hz range was significantly reduced in FXS (*M* = .17, *SD* = .09) compared to TDC (*M* = .23, *SD* = .12), as expected, *F*(1, 65) = 5.86, *p* =.018, ES = .08, OP = .67. However, the main effect of hemisphere was not significant, *F*(1, 65) = .05, *p* =.832, ES = .01, OP = .06. The observed power for the main effect of sex and hemisphere indicates the current sample is largely underpowered to detect any sex or hemisphere differences, if sex and hemisphere effects are present.

**Figure 2.**
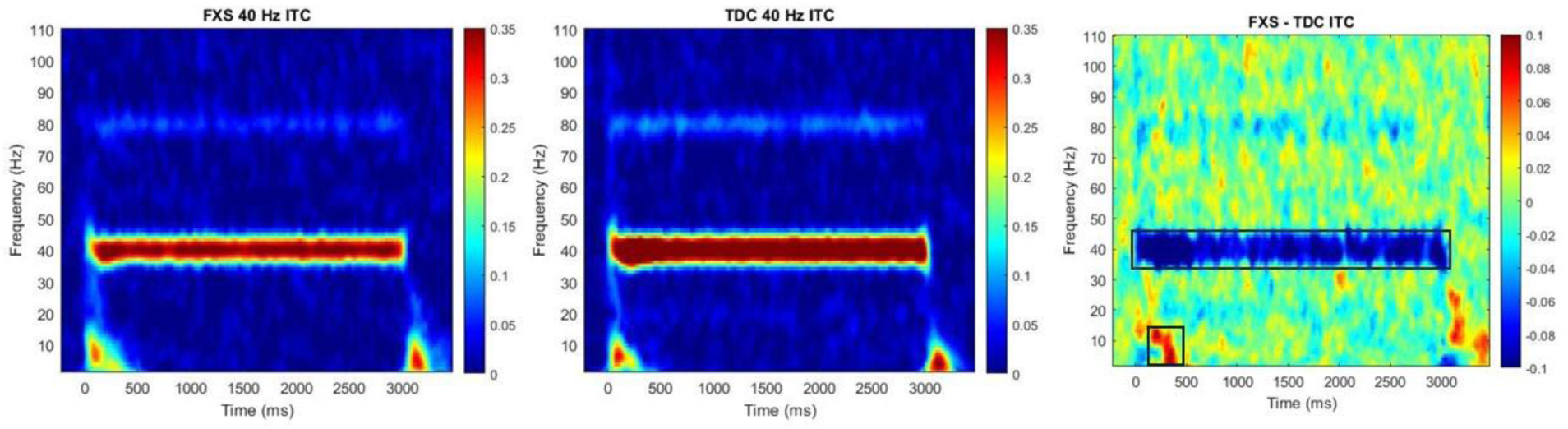
Time-frequency plots for FXS (left) and TDC (middle) with a difference plot (right) depicting group differences on ITC. Warm colors (i.e., reds and yellows) indicate increased ITC and cool colors (i.e., greens and blues) indicate reduced ITC. Black boxes indicate significant group differences.

A rmANOVA evaluated differences in 40 Hz ITC values across the duration of the stimulus between 2 components determined *a priori* (the early transient component (100 – 400 ms) and the later sustained component (650 – 3000 ms)) between groups. Results showed a significant main effect of component where ITC was elevated for the transient component (*M* = .32, *SD* = .13) compared to the sustained component (*M* = .29, *SD* = .13), *F*(1, 65) = 18.89, *p* < .001, ES = .23, OP = .99. The results also showed an expected significant main effect of group where, again, ITC was reduced for FXS (*M* = .25, *SD* = .10) compared to TDC (*M* = .33, *SD* = .13), *F*(1, 65) = 13.86, *p* < .001, ES = .18, OP = .96 (Figure 3A). The interaction between component and group was also significant which shows the main effect of component was primarily driven by TDC where TDC exhibited higher ITC during the time window for the transient component (*M* = .39, *SD* = .12) compared to the sustained component (*M* = .33, *SD* = .13) and FXS did not exhibit a significant change between the transient (*M* = .26, *SD* = .11) and the sustained components (*M* = .25, *SD* = .11), *F*(1, 65) = 9.04, *p* = .004, ES = .12, OP = .84 (Figure 3B).

**Figure 3.**
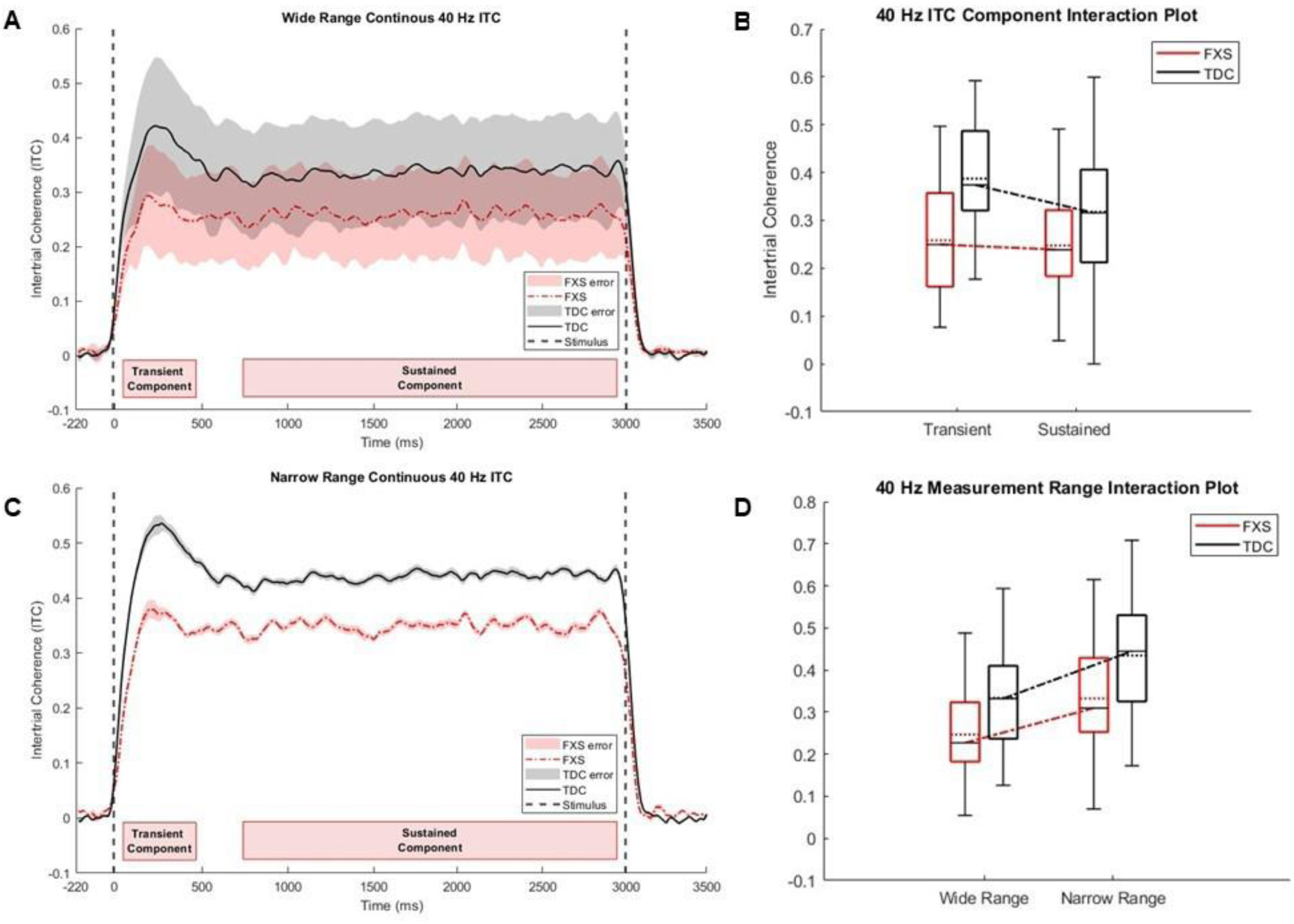
**A)** Continuous 40 Hz ITC for the wide range (35 – 45 Hz) where lines represent the mean ITC and error shading represents standard deviation point-by-point. Red boxes at the bottom represent time windows for the transient and sustained components. **B)** Box and whisker plot for the 40 Hz ITC components showing the interaction effect for component by group for the widened 40 Hz range only (shown in XA). **C)** Continuous 40 Hz ITC for the narrow range (∼39 – 41 Hz) where lines represent the mean ITC and error shading represents standard deviation point-by-point. **D)** Box and whisker plot for the 40 Hz ITC measurement range analysis showing the interaction effect for measurement window by group.

The measurement window for the 40 Hz assessment was selected *a priori* and an exploratory analysis was conducted to evaluate whether individuals with FXS show changes in neural specificity for the stimulus frequency vs. adjacent frequency ranges. ITC measurement range was narrowed from 35 – 45 Hz to 39 – 41 Hz to identify whether group differences depended on the spread of activation in the gamma range or were specific to 40 Hz oscillatory activity alone. Significance of the group difference holds where 40 Hz ITC is reduced for FXS (*M* = .34, *SD* = .13) relative to TDC (*M* = .43, *SD* = .15), *t*(65) = 2.99, *p* = .004, d = .73 but the values were different. In an exploratory rmANOVA between the *a priori* selected wider 40 Hz analysis range and the narrow range, there was a significant elevation in ITC for the narrow range (*M* = .38, *SD* = .15) compared to the wider 40 Hz ITC measure (*M* = .29, *SD* = .12), *F*(1, 65) = 906.68, *p* < .001, ES = .93, OP = 1.0 (Figure 3C). Interestingly, the interaction between 40 Hz range and group was significant where TDC exhibited increased ITC within a restricted range (*M* = .43, *SD* = .15) relative to the wider 40 Hz measure (*M* = .33, *SD* = .13) when compared to FXS who did not significantly increase in ITC between measurement ranges (narrow range: *M* = .33, *SD* = .13; wide range: *M* = .25, *SD* = .10), *F*(1, 65) = 6.14, *p* = .016, ES = .09, OP = .69 (Figure 3D).

A fourth rmANOVA tested for component differences (transient vs. sustained) for the narrow 40 Hz ITC range and showed a similar result to the rmANOVA testing the components generated from the wider 40 Hz range (Figure 3C). There was a main effect of component where ITC for the transient component (*M* = .42, *SD* = .16) was significantly elevated compared to the sustained component (*M* = .38, *SD* = .15), *F*(1, 65) = 15.14, *p* < .001, ES = .19, OP = .97. The interaction was also significant, identifying a similar dynamic to the components computed from the wider 40 Hz range where TDC exhibit a significant drop in ITC from the transient component (*M* = .50, *SD* = .14) to the sustained component (*M* = .43, *SD* = .16) and FXS does not exhibit a similar reduction in ITC indicative of an inability to mount the transient response in FXS (transient component: *M* = .34, *SD* = .14; sustain component: *M* = .33, *SD* = .13), *F*(1,65) = 8.34, *p* = .005, ES = .11, OP = .81. Of interest, standard error was markedly smaller for the narrow (Figure 3C) than the wider (Figure 3A) 40 Hz range measures, suggesting that the narrower range may more robustly discriminate groups.

A fifth, exploratory rmANOVA evaluated differences in 40 Hz ITC response across the duration of the stimulus in time-limited bins (∼100 ms) between groups which identified a significant main effect of bin (*F*(29, 37) = 7.61, *p* < .001, ES = .86, OP = 1.0) and group (*F*(1, 65) = 9.44, *p* = .003, ES = .13, OP = .86) but the interaction was not significant (*F*(29, 37) = 1.23, *p* = .275, ES = .49, OP = .76). A Bonferroni correction was applied to the pairwise comparisons between bins which identified the main effect of bin was driven by bin 1, as bin 1 was significantly different from all other bins and mostly represents both groups mounting the ASSR. The greatest bin difference was between bin 1 and bin 3 (Mean difference = -.14, SE = .01, *p* < .001) and bin 3 was significantly different from all but 8 bins, including bins 2 and 4. Bins that were not significantly different from 3 were interspersed throughout the stimulus response and generally corresponded to the peaks in ITC fluctuations (e.g., bin 13; Figure 4) indicating that bin 3, is the most extreme point of the ASSR (i.e., peaking ∼ 300 ms) and suggesting it may represent the peak of the transient component.

**Figure 4.**
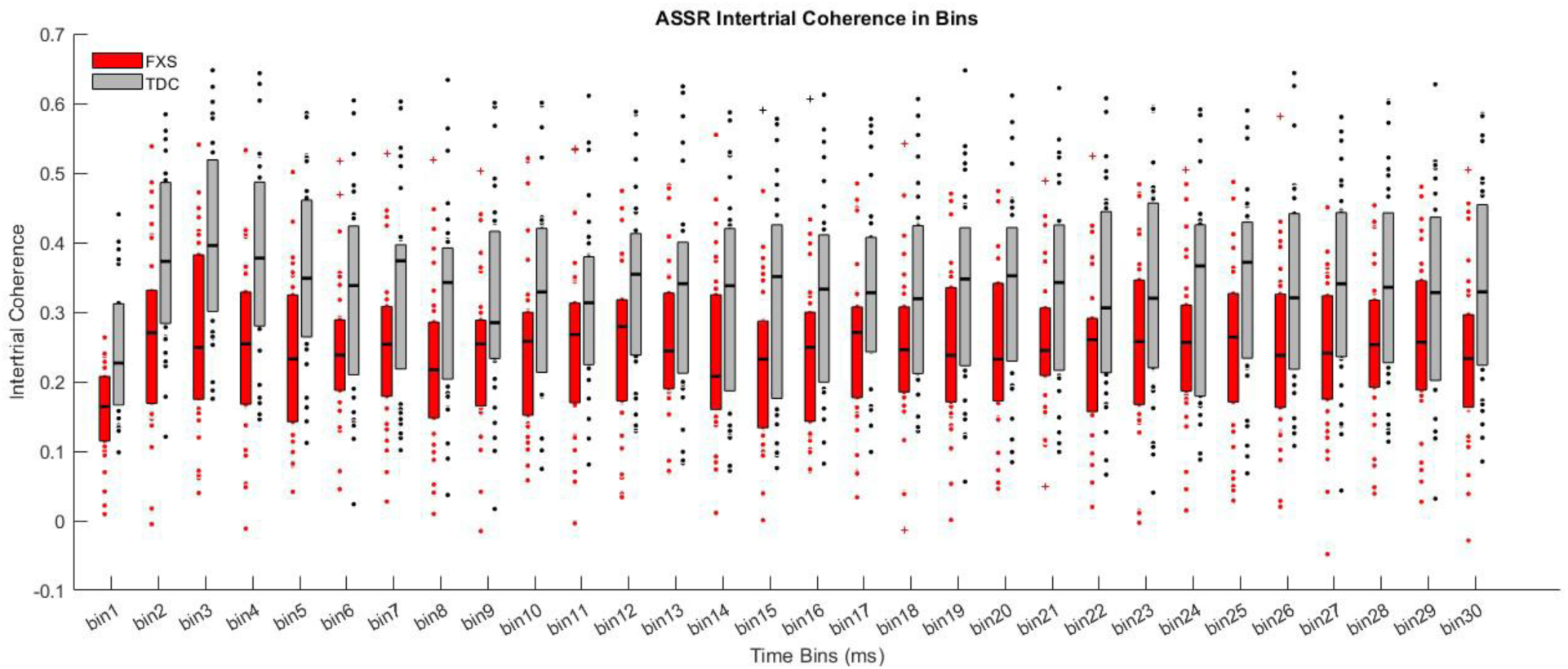
The 40 Hz ITC in time-limited bins between groups. Each bin was ∼ between 75-100 ms.

We also tested ITC in the 40 Hz range after stimulus offset between groups to evaluate whether the 40 Hz response persisted after the stimulus, and found nearly identical values in the post-stimulus response between FXS (*M* = .01, *SD* = .02) and TDC (*M* = .01, *SD* = .02), *t*(65) = .22, *p* = .827, *d* = .05.

### Additional ITC Measures

Independent Samples Mann Whitney U tests were used to evaluate low frequency onset ITC and the 80 Hz harmonic response. Stimulus onset was significantly elevated for individuals with FXS (mean rank = 39.68) compared to TDC (mean rank = 28.15), *N* = 67, *z* = 2.42, *p* = .016, r = .30. The harmonic response at 80 Hz was not significantly different between groups (FXS: mean rank = 31.79; TDC: mean rank = 36.79), *N* = 67, *z* = -.94, *p* = .347, r = .11. An independent samples t-test evaluated group differences on ITC to the offset of the stimulus and identified no significant differences between groups but the difference trended towards a significant elevation in ITC to stimulus offset for FXS (*M* = .12, *SD* = .05) compared to TDC (*M* = .10, *SD* = .05), *t*(65) = -1.78, *p* = .079, *d* = -.44 (Figure 2).

### ERSP

An independent samples t-test was used to evaluate baseline corrected ERSP in the 40 Hz range but Levene’s test for equality of variances was significant, thus Welch’s t-test was utilized. The 40 Hz power was significantly reduced for FXS (*M* = .67, *SD* = .51) compared to TDC (*M* = 1.15, *SD* = .76), *t*(56.17) = 3.02, *p* = .004, *d* = .74 (Figure 5). When evaluating the transient and sustained components for 40 Hz ERSP, we see a similar dynamic to the one identified for ITC. A rmANOVA identified a main effect of component where the transient component (*M* = 1.25, *SD* = .96) was significantly elevated relative to the sustained portion of the ASSR (*M* = 1.05, *SD* = .79), *F*(1, 65) = 14.83, *p* < .001, ES = .19, OP = .97. The main effect of group was also significant with FXS (*M* = .67, *SD* = .51) exhibiting reduced 40 Hz ERSP relative to TDC (*M* = 1.15, *SD* = .76), *F*(1, 65) = 12.35, *p* < .001, ES = .16, OP = .93. Finally, the component by group interaction was significant with TDC exhibiting a significant decrease in power from the transient component (*M* = 1.67, *SD* = 1.00) to the sustained component (*M* = 1.31, *SD* = .88) and FXS exhibiting no difference between transient (*M* = .84, *SD* = .72) and sustained (*M* = .79, *SD* = .61). The combination of reduced ITC and ERSP components suggests both temporal imprecision and overall decreased magnitude or neural engagement of the ASSR entrainment response.

**Figure 5.**
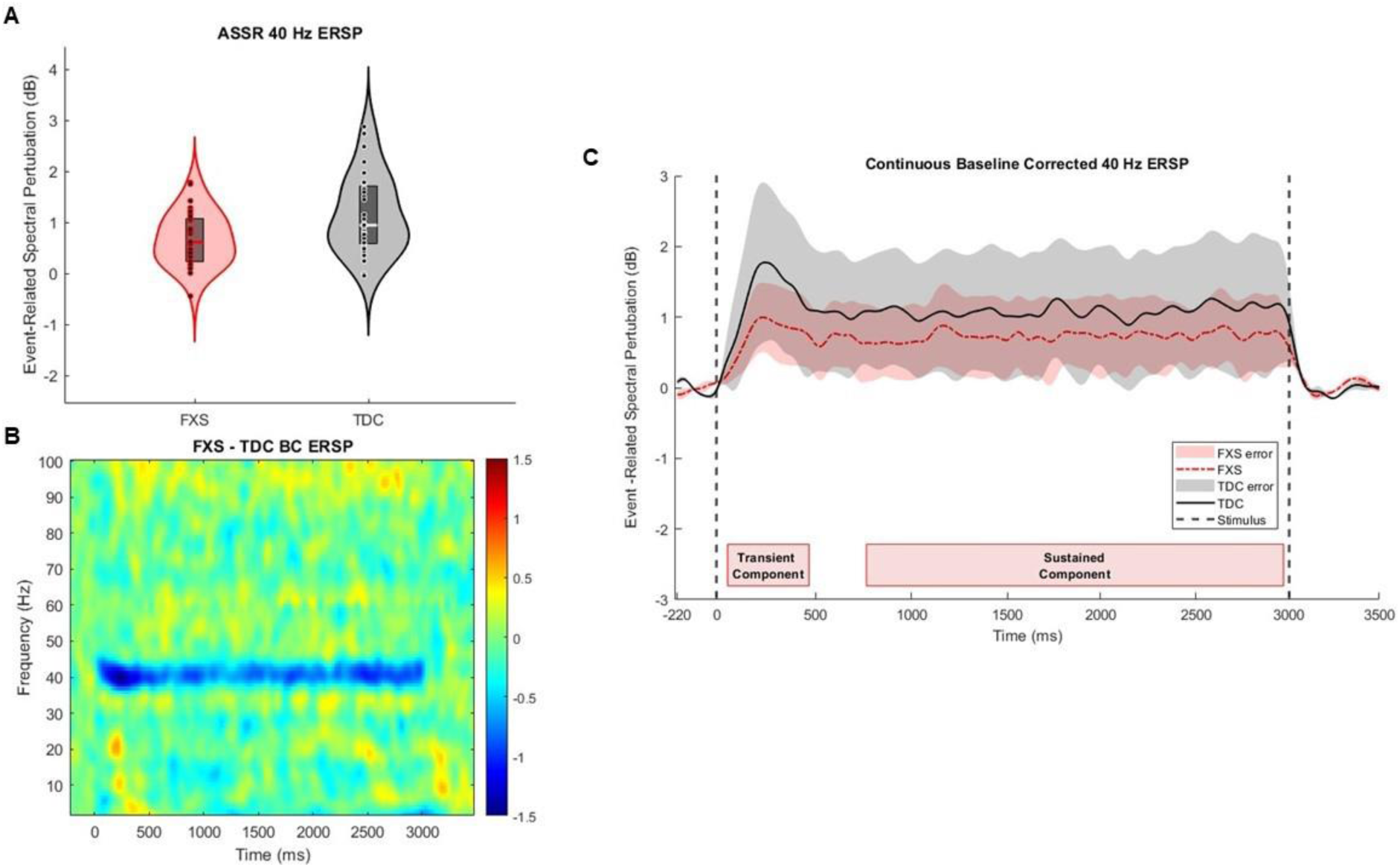
Plot depicting baseline corrected ERSP in the 40 Hz range (i.e., ASSR). **A)** Violin plot showing differences between groups. **B)** ERSP difference plot with TDC ERSP subtracted from FXS where cool colors are reductions in baseline corrected ERSP relative to TDC. **C)** Continuous ERSP line plot showing ERSP across the duration of the ASSR and indicating the window for the transient and sustained components.

Independent samples t-tests were used to test group differences on all un-baseline-corrected ERSP variables (Figure 6). Theta power was significantly elevated in FXS (*M* = 28.26, *SD* = 3.29) compared to TDC (*M* = 25.39, *SD* = 2.52), *t*(65) = -4.00, *p* < .001, *d* = -.98. Interestingly, alpha power was also significantly elevated for FXS (*M* = 24.83, *SD* = 3.32) compared to TDC (*M* = 23.08, *SD* = 3.39), *t*(65) = -2.13, *p* = .037, *d* = -.52. Finally, beta power was elevated for FXS (*M* = 18.48, *SD* = .99) relative to TDC (*M* = 16.71, *SD* = 2.35), *t*(65) = - 3.35, *p* = .001, *d* = -.82.

**Figure 6.**
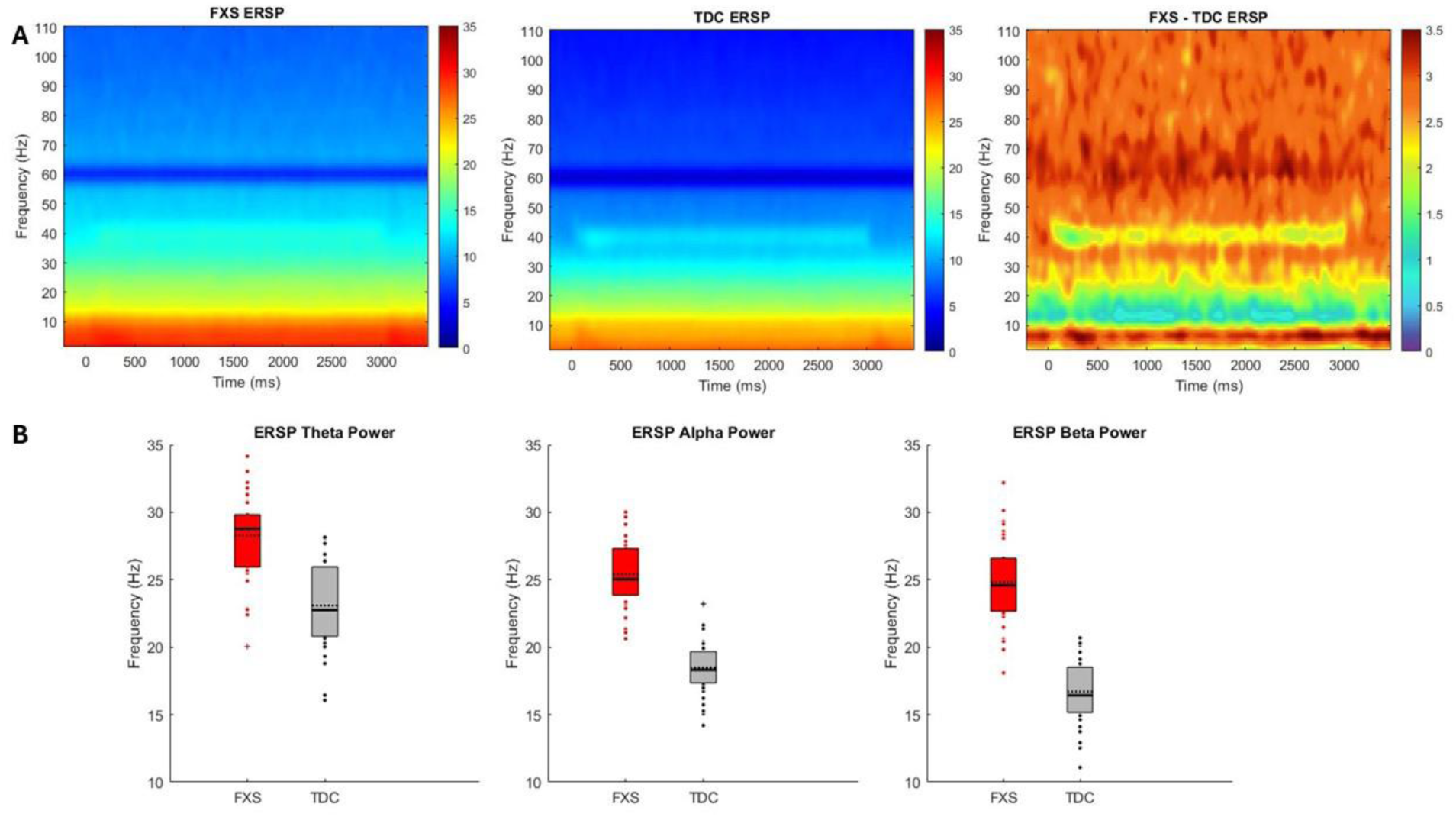
Plot depicting uncorrected ERSP results. **A)** ERSP plots for FXS (left) and TDC (middle) followed by the difference plot showing group differences. Note for the difference plot, the scale starts at 0 and all colors represent significant elevation in ERSP for FXS relative to TDC across theta, alpha, and beta frequency band ranges. **B)** Box and whisker plots for theta, alpha, and beta ERSP, respectively. The solid line on each box indicates the median and the dashed line indicates the mean.

**Figure 7.**
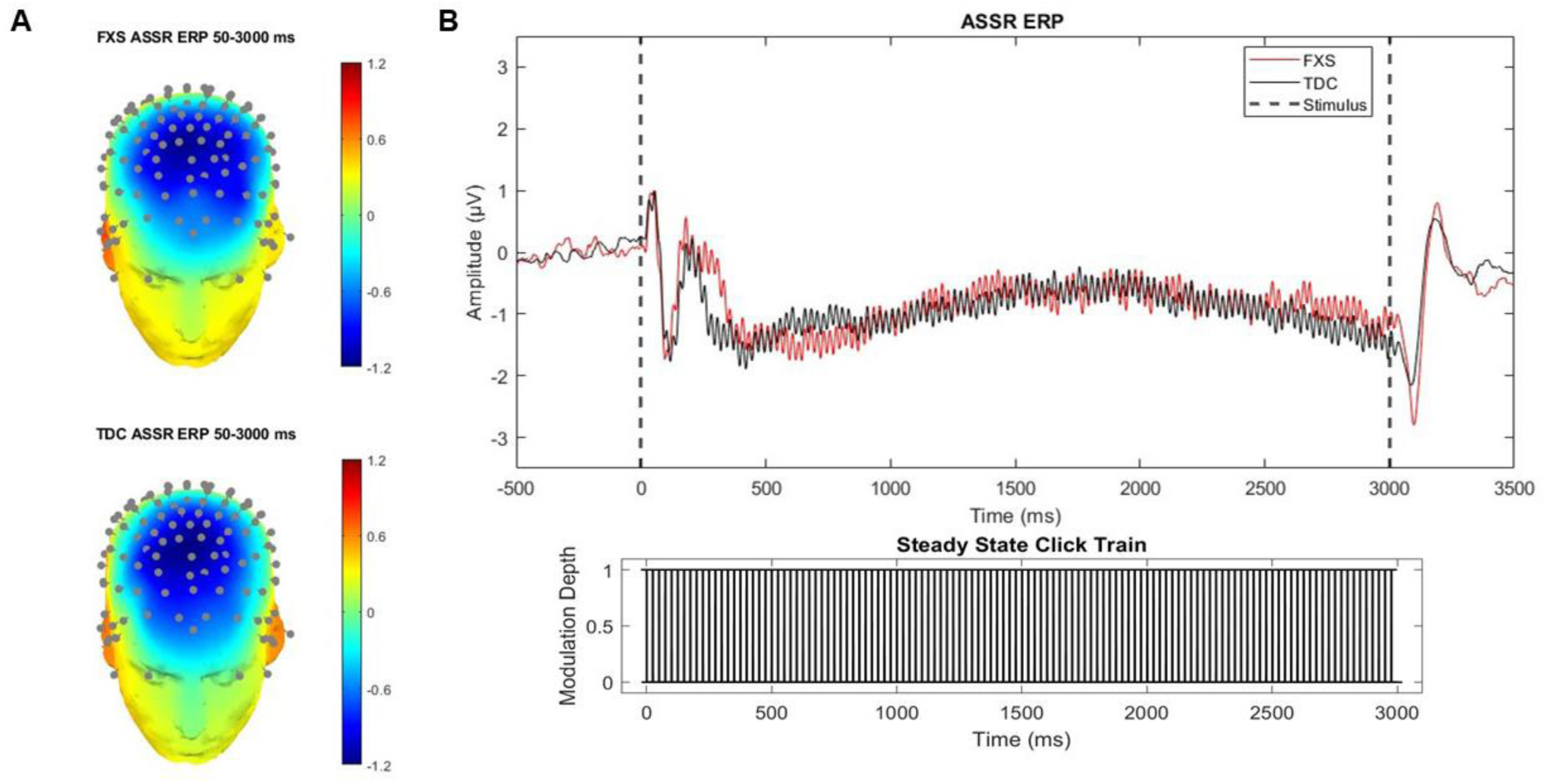
**A)** ASSR ERP scalp distribution of voltage. **B)** ASSR ERP average waveform for FXS and TDC. Note the onset P2 amplitude range is wider for FXS compared to TDC but onset P2 results are driven by the first peak

### ASSR ERP

Mann Whitney U tests were used to evaluate group differences for ASSR onset and offset ERPs. The onset P2 ERP component amplitude was significantly different between groups where individuals with FXS exhibited increased amplitudes (*M* rank = 40.03) compared to TDC (*M* rank = 27.79), z = 2.57, *N* = 67, *p* = .010, r = .32. The onset P2 latency was also significantly different between groups with FXS exhibiting faster latencies (*M* rank = 26.04) compared to TDC (*M* rank = 42.20), *z* = -3.39, *N* = 67, *p* < .001, r = -.41. The P1 and N1 ERP component amplitudes and latencies for the onset ERP were not significantly different between groups but the N1 latency trended toward an faster latency for FXS (*M* rank = 29.74) compared to TDC (*M* rank = 38.39), *z* = -1.82, *p* = .068, r = -.22.

The ERP components for the offset response were all significantly different or trended towards a significant difference between groups. The offset N1 amplitude was significantly elevated in FXS (*M* rank = 39.09) compared to TDC (*M* rank = 28.76), *z* = 2.17, *N* = 67, *p* = .030, r = .27. The offset N1 latency was significantly slower for FXS (*M* rank = 43.81) compared to TDC (*M* rank = 23.89), *z* = 4.20, *N =* 67, *p* < .001, r = .51.The offset P2 amplitude was also elevated in FXS (*M* rank = 42.59) compared to TDC (*M* rank = 25.15), *z* = 3.66, *p* < .001, r = .45. Finally, the offset P2 latency was not significantly different but did trend toward an increase in latency for FXS (*M* rank = 38.43) compared to TDC (*M* rank = 29.44), *z* = 1.89, *p* = .059, r = .23.

### Clinical Correlations

The correlation for FXS males only between log transformed peripheral FMRP and both 40 Hz ITC (rho = .036, *p* = .874) and ERSP (rho = .244, *p* = .273) were not significant. However, there was a significant correlation between FMRP and the post 40 Hz ITC (rho = .440, *p* =.041) and the N1 component amplitude to the offset (rho = -.464, *p* = .030).

Clinical correlations between EEG variables of interest and clinical variables are in supplement.

## Discussion

The present study extends previous work investigating auditory processing in humans with FXS by evaluating the ASSR. On par with previous findings, we identified reduced low gamma ITC and ERSP in FXS compared to TDC participants in response to 40 Hz steady state stimuli (Ethridge et al., 2017; 2019). The reduction of the 40 Hz ERSP and ITC are suggestive of a reduced ability to mount a low gamma response to the stimulus and reduced temporal precision of the limited activity they are able to generate, respectively. Exploratory analyses also identified a reduced ability for FXS to mobilize the early, transient component when compared to the typically developed ASSR. Our findings highlight a unique auditory dynamic in FXS where both reduced ITC and ERSP in the 40 Hz range are unchanging over the course of the ASSR which may provide unique access to the more specific cellular-level disruptions notable in *Fmr1^-/-^* mice (Jonak et al., 2024).

Decreased ERSP at 40 Hz likely represents diminished activity of neural populations responsible for generating sustained oscillatory activity in the low gamma range which may reflect the reduced synaptic maturity noted in FXS and the disruptions to the excitatory/inhibitory balance characteristic of PV+ interneuron dysfunction in FXS (Nomura et al., 2021; Salcedo-Arellano et al., 2020). The reduction in ITC, or reduced phase consistency, was also noted in rodent models of FXS and are representative of the broader frequency tuning curve in FXS auditory cortex (Jonak et al., 2024; Rotschafer & Razak, 2013). Importantly, when the measurement range was narrowed for ITC, we noted a significant interaction where the narrowed range resulted in increased ITC for TDC but not for FXS, though they moved in the same direction. While FXS are still mounting a response, the interaction suggests the ASSR is more diffuse in FXS compared to TDC which is similar to slice level findings where *Fmr1^-/-^*deficient neurons are able to fire in the gamma range but fire at less precise frequencies (Goswami et al., 2019; Lovelace et al., 2020; Rotschafer & Razak, 2013).

Factoring in the dynamics between transient and sustained components for both measures, reduced ITC and ERSP in the 40 Hz range represent an overarching reduction in the ability to mount and maintain accurate stimulus representation via neural entrainment, likely representative of both reduced GABAergic and NMDAr function (Toso et al., 2024). Specifically, the dynamics identified in the current study potentially represent reduced GABergic modulation of the ASSR and decreased balance between NMDAr and GABAergic activity where the sustained component is maintained by excitatory/inhibitory homeostasis which may reflect NMDAr hypoactivity and the lack of a transient component response is more reflective of GABAergic modulation of the ASSR (Toso et al., 2024). The present study also adds evidence in support of the conclusion that FXS is a disorder of thalamocortical disruptions given input from the thalamus, specifically the medial geniculate nucleus, is required to generate and sustain neural entrainment in the gamma range (Li et al., 2024; Pedapati et al., 2022). The transient component of the ASSR is also reflective of thalamocortical synchronization and the reduced ability to mount this response in FXS adds support to the thalamocortical dysregulation hypothesis (Franowicz & Barth 1995; Pantev et al., 1996).

Of note, a recent magnetoencephalography (MEG) study used lorazepam (GABA_a_ receptor agonist) and memantine (NMDAr antagonist) to pharmacologically challenge the ASSR in healthy, typically developed adults and found a significant elevation in percent power change, a measure comparable to ERSP, for the transient component only while participants were on lorazepam suggesting the transient component is sensitive to alterations in GABAergic function (Toso et al., 2024). Another study evaluating memantine in patients with schizophrenia and healthy controls identified an increase in both phase locking and evoked power with memantine at a higher dose (Light et al., 2017; Swerdlow et al., 2024). Additionally, a study in rats localized the ASSR to primary auditory cortex and found another NMDAr antagonist increased ASSR ITC which suggests the ASSR is sensitive to NMDAr disruptions, as well (Sullivan et al., 2015).

In addition to ERSP and ITC findings, we observed increased phase locking to the low frequency stimulus onset which is a robust neurophysiological phenotype in the FXS literature. Previously, increased onset ITC was interpreted to primarily reflect the elevated N1 amplitude, although in some auditory tasks the P2 amplitude is also elevated for FXS (Ethridge et al., 2017; 2019). However, we found that elevated onset ITC in FXS in the ASSR task additionally implicates P2 amplitude elevations.

The significant elevation for FXS in power within both uncorrected alpha and beta frequency bands represents a novel finding. Studies have identified relative power differences between FXS and TDC in the alpha range but not absolute power (Ethridge et al., 2016; Pedapati et al., 2022). Increased uncorrected alpha ERSP, which is an absolute power measure, may represent a transient fluctuation or a novel dynamic where alpha range activity is elevated by continuous stimulation at a single frequency (i.e., 40 Hz) in order to regulate the sustained high frequency activity. The elevated beta ERSP may represent a “bleeding” effect of the ASSR, where stimulation at 40 Hz results in a frequency imbalance in the beta range as a byproduct of impaired inhibitory control. Increased frequency activity in beta may be compensatory for gamma band and known low frequency disruptions in FXS (Pedapati et al., 2022). Ultimately, elevation in beta for FXS may represent similar mechanistic disruptions underlying ASSR abnormalities including NMDAr hypofunction and disrupted GABAergic signaling by means of “beat skipping” resulting from GABA decay time alterations, like seen in schizophrenia models (Metzner & Steuber, 2021; Vierling-Claassen et al., 2008). Elevated beta power has also been linked to GABAergic modulation in Tuberous Sclerosis Complex, another neurodevelopmental disorder associated with intellectual disability and autism risk (Clements et al., 2025), with NMDA antagonism in rodents (Qin et al., 2023) and with impaired cognition in schizophrenia (Donati et al., 2024). However, more evidence and refined experimentation are required to both replicate and better understand the elevations in alpha and beta ERSP before conclusions are drawn.

Finally, the analysis of the 40 Hz response in time bins, in addition to the component analysis, was an exploratory effort to address whether the ASSR was consistent throughout the stimulus duration or whether ITC changed with time. The bin analysis findings were primarily reflective of the shift from transient to sustained components where bin 3 likely represents the most extreme point of the transient component. Importantly for clinical scalability, this finding that group differences are stable throughout the sustained component of the ASSR supports the ability to shorten the stimulus, thus decreasing participant burden, particularly for individuals with sensory sensitivities, and achieve the same results.

### Limitations

While the sample was well powered to detect the primary and main exploratory analyses, it was underpowered to detect sex and hemisphere effects observed in previous studies using the auditory chirp task and rodent model studies investigating the ASSR (Norris et al., 2022; Jonak et al., 2024). Additionally, the 80 Hz harmonic was not significantly different despite seeing differences in the *Fmr1^-/-^* mice from wild type (Jonak et al., 2024). The human 80 Hz harmonic response is seemingly less robust in TDC than noted in wild type mice which represents a translational limitation (Jonak et al., 2024). Give group differences were present in the *Fmr1^-/-^* mice, the current study may also be underpowered to detect differences in the harmonic response.

## Conclusions

The current results provide increased evidence in support of the conclusion that FXS is a disorder of impaired thalamocortical circuitry and increased cortical temporal variability/imprecision. The ASSR highlights novel disruptions via analysis of the transient and sustained components, which may provide increased access to disruptions at the cellular level making the steady state paradigm ideal for translational use. The ASSR also replicates many rodent findings with the same stimulus (Jonak et al., 2024) and previously documented auditory processing differences in FXS identified utilizing different auditory paradigms (Ethridge et al., 2017; 2019). Both pharmacological intervention and back-translation of the component dynamics in mice may inform which circuitry disruptions contribute to overall reduction in low gamma neural entrainment and the inability to mount the transient response in the 40 Hz range.

## Data Availability

The dataset used and/or analyzed during the current study are available from Lauren Ethridge on reasonable request.

## Acknowledgements

We thank the participants with FXS and their families for their participation and continued support of our program of research. We additionally would like to acknowledge the passing of the late Dr. John Sweeney, who was a mentor to many of the authors and contributed to the conceptualization of the reported work.

## List of Author Contributions

JEN led all EEG data pre-processing/artifact rejection, conducted all analyses, and drafted the manuscript; LAD consulted on data analyses and co-led data collection and management; WSM managed clinical and FMRP variable curation and aided with clinical data interpretation; LMS led all EEG and clinical data collection and contributed to both FMRP and clinical data analysis and interpretation; CG, SP, and BH generated FMRP variables and led FMRP data management; EVP assisted with all EEG data collection; CAE supervised all aspects of study design, funding, and participant recruitment; LEE supervised all aspects of EEG experimental design, data analysis, and manuscript preparation. All authors contributed significantly to manuscript preparation and editing. All authors approve of the final draft.

