## Supplemental Materials for "Auditory steady-state response deficits in Fragile X Syndrome implicate deficits in stimulus representation maintenance and GABAergic modulation"

**Supplement**

**Pharmaceutical Information**

In total, three individuals were on anticonvulsants the day of EEG recording, two had taken an anticonvulsant at least 24 hours prior to the EEG recording, four were on atypical antipsychotics, eleven were on antidepressants with nine on selective serotonin reuptake inhibitors (SSRIs) and two on a serotonin-norepinephrine reuptake inhibitor (SNRI), and two on anti-anxiolytics. Six individuals were on both SSRIs and atypical antipsychotics with one individual taking two different atypical antipsychotics simultaneously. Finally, two individuals listed above were taking muscle relaxants, one was on an anti-tremor medication, four were on a stimulant, and one was taking a cognition boosting, non-stimulant ADHD medication.

**Supplemental Table 1.**

*EEG Outcomes by Anti-Convulsant Use Status*

| | **Participant AC Use Status** | | | | | |
|---|---|---|---|---|---|---|
| | **On day-of-AC (*N* = 3)** | | **No day-of-AC FXS (*N* = 31)** | | **No day-of-AC (All) (*N* = 64)** | |
| **EEG Variables** | **M** | **SD** | **M** | **SD** | **M** | **SD** |
| **ITC** | | | | | | |
| Onset ITC | .12 | .02 | .15 | .07 | .12 | .06 |
| Offset ITC | .15 | .03 | .12 | .05 | .11 | .05 |
| 40 Hz ITC | .26 | .13 | .25 | .10 | .29 | .12 |
| Narrow 40 Hz ITC | .34 | .16 | .33 | .13 | .38 | .15 |
| Post 40 Hz | .02 | .02 | .01 | .02 | .01 | .02 |
| 80 Hz ITC | .06 | .07 | .03 | .02 | .04 | .04 |
| 40 Hz ITC Transient component | .27 | .11 | .26 | .12 | .32 | .14 |
| 40 Hz ITC Sustained component | .26 | .13 | .25 | .11 | .28 | .13 |
| Narrow 40 Hz ITC Transient component | .37 | .16 | .34 | .14 | .42 | .16 |
| Narrow 40 Hz ITC Sustained component | .35 | .17 | .33 | .13 | .38 | .15 |
| **ERSP** | | | | | | |
| Theta ERSP | 26.62 | 2.24 | 28.42 | 3.35 | 26.86 | 3.30 |
| Alpha ERSP | 22.41 | 3.22 | 25.07 | 3.29 | 24.04 | 3.46 |
| Beta ERSP | 19.48 | .73 | 18.39 | 2.06 | 17.52 | 2.35 |
| BC 40 Hz ERSP | .65 | .50 | .68 | .50 | .91 | .69 |
| 40 Hz ERSP Transient Component | .64 | .87 | .86 | .72 | 1.25 | .96 |
| 40 Hz ERSP Sustained Component | .81 | .65 | .79 | .62 | 1.05 | .79 |
| **ERP** | | | | | | |
| Onset P1 Amplitude | .06 | .01 | .04 | .02 | .04 | .02 |

| | | | | | | |
|---|---|---|---|---|---|---|
| Onset P1 Latency | 46.33 | 5.77 | 46.45 | 6.28 | 46.53 | 6.06 |
| Onset N1 Amplitude | .06 | .02 | .13 | .08 | .12 | .07 |
| Onset N1 Latency | 100.33 | 12.50 | 112.13 | 9.34 | 112.75 | 10.20 |
| Onset P2 Amplitude | 1.14 | .60 | 1.35 | .69 | 1.15 | .63 |
| Onset P2 Latency | 240.00 | 16.46 | 1.35 | 23.29 | 244.44 | 24.57 |
| Offset N1 Amplitude | .09 | .05 | .18 | .09 | .15 | .08 |
| Offset N1 Latency | 3113.67 | 8.51 | 3105.29 | 8.50 | 3101.95 | 8.32 |
| Offset P2 Amplitude | .13 | .01 | .125 | .07 | .10 | .061 |
| Offset P2 Latency | 3209.33 | 25.66 | 3199.35 | 13.41 | 3196.23 | 13.67 |

*Note.* Bolded values were statistically significant. BC = baseline corrected. Blank = NS. *$p < 0.05$, **$p < 0.01$, ***$p < .001$.

**Exploratory Correlations – EEG**

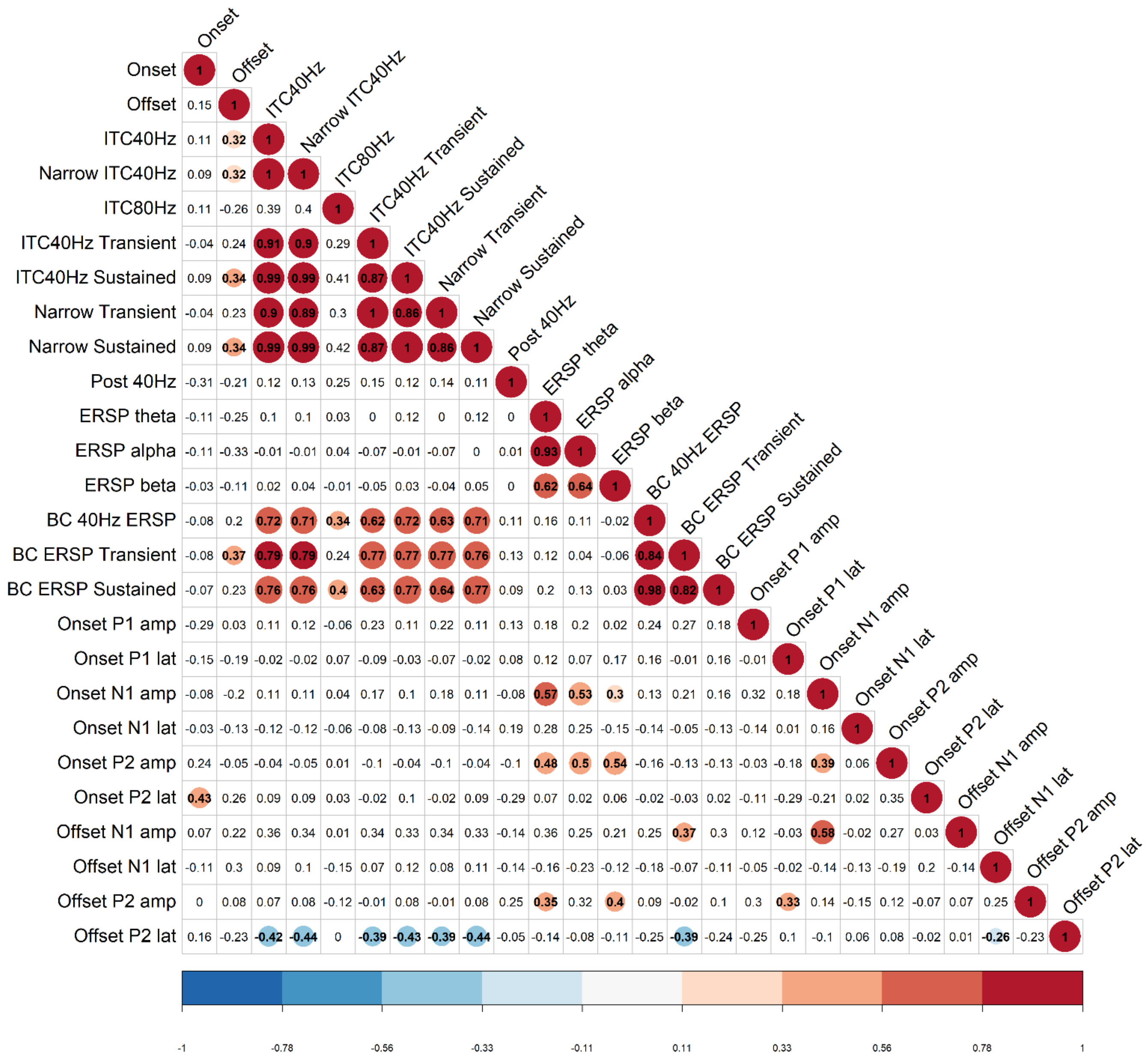


**Supplemental Figure 1.** Exploratory correlation values between EEG variables of interest. Bolded values with circles were statistically significant, and all non-significant correlations have no circle. Color coding corresponds to p-values with cooler colors representing inverse correlations, and all warmer colors representing direct correlations. Narrow = narrowed 40 Hz range; BC = baseline corrected; amp = amplitude; lat = latency.

**Exploratory Correlations – Clinical Measures**

Correlations between EEG variables interest and A/ASP, SB-5, CSP, ABC, and KiTAP measures are listed in Supplemental Tables 2-6. The SCQ, WJ-III, Vineland, and ADAMS were all not correlated with any of the EEG variables of interest for FXS and are not reported for the full FXS sample.

***Supplemental Table 2.***

*Correlations Between EEG Variables of Interest and SB-5 IQ Variables*

| EEG Variable | Stanford-Binet 5th Edition Nonverbal IQ (N = 32) | Stanford-Binet 5th Edition Verbal IQ (N = 31) | Stanford-Binet 5th Edition Abbreviated IQ (N = 33) |
|---|---|---|---|
| **ITC** | | | |
| Onset | | | |
| Offset | | | |
| 40 Hz | | | |
| Narrow 40 Hz | | | |
| Transient Component | | | |
| Sustained Component | | | |
| 80 Hz (harmonic) | | | |
| **ERSP** | | | |
| Alpha | | | |
| Theta | | | |
| Beta | **-.460**** | **-.371*** | **-.447**** |
| Baseline corrected 40 Hz | .323 (.71) | **.355*** | .310 (.079) |
| **ERP - Onset** | | | |
| P1 Amplitude | | | |
| N1 Amplitude | | | |
| P2 Amplitude | | | |
| P1 Latency | | | |
| N1 Latency | | | |
| P2 Latency | | | |
| **ERP - Offset** | | | |
| N1 Amplitude | | | |
| P2 Amplitude | | | |
| N1 Latency | -.336 (.06) | **-.510**** | |
| P2 Latency | | | |

*Note.* Bolded values were statistically significant. All SB-5 variables are deviation IQ. Blank = NS. **p* < 0.05, ***p* < 0.01, ****p* < .001.

***Supplemental Table 3.***

*Correlations Between EEG Variables of Interest and A/ASP Subscale Scores*

| EEG Variable | Adolescent and Adult Sensory Profile Variables | | | | |
|---|---|---|---|---|---|
| | Low Registration | Sensation Seeking | Sensory Sensitivity | Sensory Avoidance | Auditory |
| **ITC** | | | | | |
| Onset | | | | | |
| Offset | | | | | |
| 40 Hz | .352 (.078) | | | | |
| Narrow 40 Hz | .343 (.086) | | | .337 (.092) | |
| Transient Component | | | | | |
| Sustained Component | .346 (.084) | | | | |
| 80 Hz (harmonic) | | | | | |
| **ERSP** | | | | | |
| UC Alpha | | | | | |
| UC Theta | | | | | |
| UC Beta | | | | | |
| BC 40 Hz | **.450*** | | **.389*** | **.448*** | |
| BC 40 Hz Transient | | | | | |
| BC 40 Hz Sustained | **.452*** | | .358 (.072) | **.443*** | |
| **ERP - Onset** | | | | | |
| P1 Amplitude | | | | | |
| N1 Amplitude | | | | | |
| P2 Amplitude | | | | | |
| P1 Latency | **.444*** | | | | |
| N1 Latency | | | | | |
| P2 Latency | | | | | |
| **ERP - Offset** | | | | | |
| N1 Amplitude | | | | | |
| P2 Amplitude | **.391*** | | | | |
| N1 Latency | | | | | |
| P2 Latency | | | | | |

*Note.* Bolded values were statistically significant. Abb. BC = baseline corrected; UC = non-baseline corrected ERSP; LR = low registration; Sen = sensation; SS = sensory sensitivity; Blank = NS. *$p < 0.05$, **$p < 0.01$, ***$p < .001$.

***Supplemental Table 4.***

*Correlations Between EEG Variables of Interest and CSP Subscale Scores*

| | | | **Child Sensory Profile (CSP) Subscales** | | |
|---|---|---|---|---|---|
| **EEG Variable** | Seek | Avoid | Sensitivity | Bystander | Auditory |
| **ITC** | | | | | |
| Onset | | | | | |
| Offset | | | .327 (.090) | | |
| 40 Hz | | | | | |
| Narrow | | | | | |
| 40 Hz | | | | | |
| Transient Component | | | .341 (.075) | | |
| Sustained Component | | | | | |
| 80 Hz (harmonic) | | | | | |
| **ERSP** | | | | | |
| UC Alpha | | | | | |
| UC Theta | | | | | |
| UC Beta | | | | | |
| BC 40 Hz | | .366 (.060) | | **.445*** | |
| BC 40 Hz Transient | | .352 (.072) | | .373 (.096) | |
| BC 40 Hz Sustained | | .364 (.062) | | **.491*** | |
| **ERP - Onset** | | | | | |
| P1 Amplitude | | | | | |
| N1 Amplitude | | | | .383 (.086) | .323 (.093) |
| P2 Amplitude | | | | | |
| P1 Latency | .326 (.090**)** | .362 (.064) | | | |
| N1 Latency | | | | | |
| P2 Latency | | | | | |
| **ERP - Offset** | | | | | |
| N1 Amplitude | | | | | |
| P2 Amplitude | | | | | |
| N1 Latency | | | | | |
| P2 Latency | | | | | |

*Note.* Bolded values were statistically significant. Abb. BC = baseline corrected; UC = non-baseline corrected ERSP; LR = low registration; Sen = sensation; SS = sensory sensitivity; Blank = NS. *$p < 0.05$, **$p < 0.01$, ***$p < .001$.

***Supplemental Table 5.***

*Correlations Between EEG Variables of Interest and ABC Subscales*

| | ABC Subscales | | | | | |
|---|---|---|---|---|---|---|
| **EEG Variable** | Irritability | Lethargy | Stereotypy | Hyperactivity | Inappropriate Speech | Social Avoidance |
| **ITC** | | | | | | |
| Onset | | | | **-.560**** | | |
| Offset | | | | | | |
| 40 Hz Narrow | | | | | | |
| 40 Hz Transient Component | | | | | | |
| Sustained Component | | | | | | |
| 80 Hz (harmonic) | | | | | | |
| **ERSP** | | | | | | |
| Alpha | | | | | | |
| Theta | | | | | | |
| Beta | | | | | | |
| Baseline corrected 40 Hz | | | | | | |
| **ERP - Onset** | | | | | | |
| P1 Amplitude | **-.463*** | | -.359 (.056) | | **-.466*** | **-.433*** |
| N1 Amplitude | | | | | | |
| P2 Amplitude | | | | | | |
| P1 Latency | | **.396*** | | | | |
| N1 Latency | | | | | | |
| P2 Latency | | | | | | |
| **ERP - Offset** | | | | | | |
| N1 Amplitude | | | | | | |
| P2 Amplitude | | | | | | |
| N1 Latency | | | **.371*** | .364 (.057) | **.404*** | .324 (.086) |
| P2 Latency | | | | | | |

*Note.* Bolded values were statistically significant. Blank = NS. *$p < 0.05$, **$p < 0.01$, ***$p < .001$.

***Supplemental Table 6.***

*Correlations Between EEG Variables of Interest and KiTAP Behavioral Variables*

| | KiTAP Variables | | | | | | | | |
|---|---|---|---|---|---|---|---|---|---|
| **EEG Variable** | Alert Median | Alert Correct | Distract Correct | Distract Error | No Distract Correct | No Distract Error | Go-NoGo Median | Go-NoGo Correct | Go-NoGo Errors |
| **ITC** | | | | | | | | | |
| Onset | | | | | -.385 (.057) | | | | |
| Offset | | **-.481*** | | | -.340 (.097) | | **.430*** | | |
| 40 Hz | | | | | | | | | **-.411*** |
| Narrow 40 Hz | | | | | | | | | **-.407*** |
| Transient Component | | | | | | | | | **-.418*** |
| Sustained Component | | | | | | | | | -.389 (.054) |
| 80 Hz (harmonic) | | | | | | | | .392 (.053) | **-.475** |
| **ERSP** | | | | | | | | | |
| UC Alpha | | | | | | | | | |
| UC Theta | | | | | | | | | |
| UC Beta | | -.364 (.074) | | | | | | | |
| BC 40 Hz | | | | | | .388 (.056) | | .359 (.078) | |
| BC 40 Hz Transient | | | | | | | | | |
| BC 40 Hz Sustained | | | | | | .344 (.092) | | .347 (.089) | |
| **ERP - Onset** | | | | | | | | | |
| P1 Amplitude | | | | | | | | | |
| N1 Amplitude | | | | | | | | | |
| P2 Amplitude | | | | | | | | | -.380 (.061) |
| P1 Latency | | | | | -.355 (.082) | | | | |
| N1 Latency | | | | | | | | | |
| P2 Latency | | -.367 (.071) | | | | | | | |
| **ERP - Offset** | | | | | | | | | |
| N1 Amplitude | | | | | | | | | |
| P2 Amplitude | | | | | **-.507**** | | | | |
| N1 Latency | | | | | | | .346 (.090) | | |
| P2 Latency | **.576**** | | **-.509**** | | | | **.478*** | | |

*Note.* Bolded values were statistically significant. Abb. BC = baseline corrected; UC = non-baseline corrected ERSP. Blank = NS. *$p < 0.05$, **$p < 0.01$, ***$p < .001$.

**Clinical Correlation Discussion**

The correlations between EEG variables and both the SB-5 and the A/ASP are most relevant to clinical concerns regarding auditory processing and cognitive sequelae. We regard the CSP correlations as more confirmatory for correlations with the A/ASP, given the A/ASP is more commonly used to evaluate sensory experiences in adolescents and adults with FXS.

We identified inverse correlations between non-corrected beta ERSP and deviation verbal, nonverbal, and abbreviated IQ variables. Elevated non-corrected beta ERSP was interpreted to indicate a potential “bleeding” effect of consistent stimulation at 40 Hz resulting from an imbalance in inhibitory control and the imprecision of firing frequency. Given the potential mechanistic disruptions to GABAergic modulation, the inverse correlations between beta and IQ may represent a potential disruption to the stream of information flow with downstream consequences for global cognitive performance. However, we strongly reiterate the need for replication and back translation to rodent model to better understand and definitively comment on mechanistic implications of elevated beta ERSP in FXS. Additionally, deviation verbal IQ was positively correlated with baseline corrected 40 Hz ERSP and the other IQ measures (i.e., non-verbal and abbreviated) trended toward a positive correlation with 40 Hz ERSP. As baseline corrected ERSP for the ASSR is reduced in FXS and representative of the ability to mount a response to the 40 Hz stimulus, the correlation suggests that an increased ability to generate a respond to the steady state stimulus is related to global cognitive ability which may align with findings in schizophrenia where elevated frontal beta is associated with worse cognitive performance on a goal-directed working memory task (Donati et al., 2024).

The sensory profile correlations (i.e., A/ASP and CSP) identified increased ASSR values as associated with worse sensory outcomes. This finding is counterintuitive in relation to previous findings in FXS using the auditory chirp (Ethridge et al., 2019). Contradictory findings on the relationship between ASSR and chirp ITC suggest that care should be taken when assessing therapeutic benefit of increasing ASSR in FXS, in that interventions that increase cognition and ASSR may result in more sensory issues for patients with FXS, reflecting a tradeoff between sensory and cognitive systems in FXS. Alternatively, the present correlations between EEG variables and sensory profile subscales may reflect a more complex relationship where increased sensory sensitivities are driving increased sensory vigilance and resulting in increased ASSR measures as a byproduct of attention-related sensory gating changes or general aversion to the auditory stimulation (Manting et al., 2020). Specifically, novelty detection is known to impact ASSR metrics where randomization of the ITI may result in increased responses but specifically in those primed to respond due to elevated baseline sensory sensitivities (Sugiyama et al., 2024). Again, replication and refined experimentation will increase the ability to interpret the present results and may help tease apart the correlations between EEG variables and the sensory profile.
